# Self-reported taste and smell disorders in patients with COVID-19:distinct features in China

**DOI:** 10.1101/2020.06.12.20128298

**Authors:** Jia Song, Yi-ke Deng, Hai Wang, Zhi-Chao Wang, Bo Liao, Jin Ma, Chao He, Li Pan, Yang Liu, Isam Alobid, De-Yun Wang, Ming Zeng, Joaquim Mullol, Zheng Liu

## Abstract

**Objectives:** We aimed to explore the frequencies of nasal symptoms in patients with COVID-19, including loss of smell and taste, as well as their presentation as the first symptom of the disease and their association with the severity of COVID-19.

**Methods:** In this retrospective study, 1,206 laboratory-confirmed COVID-19 patients were included and followed-up by telephone call one month after discharged from Tongji Hospital, Wuhan. Demographic data, laboratory values, comorbidities, symptoms, and numerical rating scale scores (0-10) of nasal symptoms were extracted from the hospital medical records, and confirmed or reevaluated by the telephone follow-up.

**Results:** From COVID-19 patients (N = 1,172) completing follow-up, 199 (17%) subjects had severe COVID-19 and 342 (29.2%) reported nasal symptoms. The most common nasal symptom was loss of taste (20.6%, median score = 6), while 11.4% had loss of smell (median score = 5). The incidence of nasal symptom including loss of smell and loss of taste as the first onset symptom was <1% in COVID-19 patients. Loss of smell or taste scores showed no correlation with the scores of other nasal symptoms. Loss of taste scores, but not loss of smell scores, were significantly increased in severe vs. non-severe COVID-19 patients. Interleukin (IL)-6 and lactose dehydrogenase (LDH) serum levels positively correlated with loss of taste scores. About 80% of COVID-19 patients recovered from smell and taste dysfunction in 2 weeks.

**Conclusion:** In the Wuhan COVID-19 cohort, only 1 out of 10 hospital admitted patients had loss of smell while 1 out 5 reported loss of taste which was associated to severity of COVID-19. Most patients recovered smell and taste dysfunctions in 2 weeks.

## Introduction

Last December 2019, a cluster of viral pneumonia cases identified as coronavirus disease 2019 (COVID-19), was reported in Wuhan, China. Subsequently, severe acute respiratory syndrome coronavirus 2 (SARS-CoV-2) has been identified to be the pathogenic cause of COVID-19 [1-3]. This newly recognized illness has spread rapidly throughout Wuhan (Hubei province) to allover the world [4-6]. The diagnosis of the COVID-19 is based on clinical manifestations, contact history report, chest computed tomography (CT) and positive result of nucleic acid test [7-9]. Due to the uncertainty of contact history and the rush to hospitals that could run out of the essential equipment for test during the world-wide pandemic, efforts to identify additional diagnostic or prognostic symptoms of COVID-19 has significant value in mitigating transmission.

The clinical spectrum of COVID-19 ranges from asymptomatic to severe ill cases [1, 2, 6, 7, 10-12]. The most common symptom of COVID-19 is fever, and other common systemic symptoms include dyspnea, cough, nausea and vomiting, etc [6, 7, 10]. In addition, olfactory and taste disorder have been recently noted in patients with COVID-19. An early study conducted in Italy reported 33.9% of hospitalized COVID-19 patients shown at least one taste or olfactory disorders [13]. Later studies in Europe reported that about 75%-85% COVID-19 patients had olfactory dysfunction and about 70%-88% patients had gustatory dysfunctions [14]. Very recently, similar prevalence of smell and taste disorder has been found in COVID-19 patients in USA [15]. It is indicated that smell or taste change may be a strong predictor for a COVID-19 positive test result and may serve as an early alerting symptom for COVID-19 [16]. However, an early study based on analyzing electronic medical records of 214 patients with COVID-19 in Wuhan, China reported that the ratio of patients complaining of loss of taste and loss of smell was only 5.6% and 5.1%, respectively [17], significantly lower than those reported in Europe and USA. One potential reason for the low ratios in China may be related to incomplete medical records of COVID-19 patients under actual emergency situation, which underestimated the incidences of upper airway tract manifestations. Nevertheless, it is also possible that there are different responses to SARS-CoV-2 infection in people with distinct ethnic/culture background. Therefore, more accurate evaluation of upper airway tract manifestations in COVID-19 patients should be conducted to figure out the clinical importance of smell and taste dysfunction in the early diagnosis of COVID-19 for Chinese and to elucidate whether there are different clinical manifestations between patients with distinct ethnic/culture background. Moreover, several important questions remain to be answered. How severe are the upper airway tract symptoms in patients with COVID-19? Is there any correlation between olfactory and taste disorders and other nasal symptoms such as nasal obstruction? Will the severity of olfactory or taste disorder be associated with the severity of COVID-19? Will there be a full recovery of olfactory or taste disorder and how long it will take?

In this retrospective study, we investigated the COVID-19 patients discharged from Tongji Hospital, the largest designated hospital to treat patients with COVID-19, in Wuhan. Integrating medical record analysis and reevaluation of upper airway symptoms by the telephone follow-up, we aimed to explore the frequencies of nasal symptoms in patients with COVID-19, including loss of smell and taste, as well as their presentation as the first symptom of the disease and their association with the severity of COVID-19.

## Methods

### Study participants

A single center, retrospective cohort study was conducted in Wuhan, China, where COVID-19 initially outbroke. We obtained the electronic medical records for discharged COVID-19 patients between January 27, 2020 and March 10, 2020, who initially admitted to the Tongji Hospital. The diagnosis was made on the basis of guidance for diagnosis and management of COVID-19 released by WHO [3]. A laboratory-confirmed case of COVID-19 was defined as having positive result on real-time reverse-transcriptase-polymerase-chain-reaction assay of nasal and pharyngeal swab specimens. Only laboratory-confirmed cases were included in the study. All patients were followed up by telephone on the 30th (±2) day after discharge. The study was approved by Tongji Hospital Research Ethics Committee.

### Clinical characteristic, laboratory assessment, and telephone follow-up

The degree of severity of COVID-19 was defined as severe and non-severe at the time of admission using the American Thoracic Society guidelines for community-acquired pneumonia [18]. The information of demographic characteristics, systemic major symptoms, and major commodities related to COVID-19 were extracted from electronic medical records. In addition, the results of laboratory assessments on admission were also collected from electronic medical records. All laboratory testing was performed according to the clinical care needs of the patients. Laboratory assessments consisted of a complete blood routine, blood biochemistry, coagulation function, infection biomarkers and immune function. All data were entered into a computerized database and cross-checked.

The airway comorbidities and nasal symptoms, the date of symptomatic onset, and numerical rating scale scores and duration days of symptoms were obtained based on the hospital medical records and were confirmed and reevaluated by the telephone follow-up. The severity of upper respiratory tract symptoms was scored by patients on a numerical rating scale of 0-10, with 0 being “no complaint whatsoever” and 10 being “the worst imaginable complaint” [19, 20]. Six major symptoms of upper respiratory tract were focused on: nasal obstruction, rhinorrhea, nasal itching, sneezing, loss of smell, and loss of taste. The severity of loss of smell and loss of taste was defined as follows: Mild = score 0-3; Moderate= score 4-7; Severe = score 8-10 [19, 20]. The difference between the symptom of loss of smell and loss of taste was explained to the patients very carefully during telephone follow-up according to previous studies [21, 22]. The detailed questionnaire showed in this article’s Online Supplement.

### Statistical Analysis

For continuous variables, Mann-Whitney *U* 2-tailed test was used for between-group comparison. Chi-square test was applied to compare the difference in proportions between groups. Spearman test was used for correlation analysis. Difference was considered to be statistically significant if a *P* value was less than 0.05. These statistical analyses were performed by using an IBM SPSS 22.0 package (SPSS Inc, Chicago, IL).

## Results

### Demographic and clinical characteristics

Totally 1,206 patients were enrolled and 1,172 of them completed questionnaires. The follow-up rate was 97.2%. Reasons of the lost cases included: refusal to answer questions for personal reasons (n = 23); unable to provide accurate information (n = 6); phone calls were not answered (n = 5). The demographic and clinical characteristics of 1,172 patients are shown in Table 1. The median age was 61 years (IQR, 48-68), and 577 (49.2%) were men. The ratio of severe cases was 17% (199/1172).

**Table 1.** Demographic and clinical characteristics of 1,172 etiologically confirmed patients.

| Characteristics | Total patients | Non-severe | Severe | *P* value |
| --- | --- | --- | --- | --- |
| Subject, N (%) | 1172 (100) | 973 (83.0) | 199 (17.0) | - |
| Gender, male, N (%) | 577 (49.2) | 480 (49.3) | 97 (48.7) | 0.94 |
| Age, years, median (IQR) | 61 (48, 68) | 60 (46, 68) | 64 (53, 70) | **<0.01** |
| Systemic signs and symptoms<br>N (%) | - | - | - | - |
| Fever | 921 (78.6) | 758 (77.9) | 163 (81.9) | 0.21 |
| Cough | 767 (65.4) | 625 (63.2) | 142 (71.4) | 0.06 |
| Myalgia | 172 (14.7) | 136 (14.0) | 36 (18.1) | 0.13 |
| Fatigue | 285 (24.3) | 226 (23.2) | 59 (29.7) | 0.05 |
| Anorexia | 274 (23.4) | 214 (22.0) | 60 (30.2) | **0.01** |
| Confusion | 6 (0.5) | 3 (0.3) | 3 (1.5) | **0.03** |
| Dizziness | 52 (4.4) | 41 (4.2) | 11 (5.5) | 0.41 |
| Any airway comorbidity, N (%) | 203 (17.3) | 179 (18.4) | 24 (12.1) | **0.03** |
| AR | 115 (9.8) | 99 (10.2) | 16 (8.0) | 0.43 |
| CRS | 72 (6.1) | 64 (6.6) | 8 (4.5) | 0.34 |
| Asthma | 29 (2.5) | 25 (2.6) | 4 (2.0) | 0.81 |
| COPD | 10 (0.9) | 10 (1.0) | 0 (0) | 0.23 |
| Any nasal symptom, N (%) | 342 (29.2) | 275 (28.3) | 67 (33.7) | 0.15 |
| Nasal obstruction, N (%) | 101 (8.6) | 82 (8.4) | 19 (9.5) | 0.58 |
| Score, median (IQR) | 3 (2, 4) | 3 (2, 4) | 3 (2, 5) | 0.88 |
| As first symptom, N (%) | 2 (0.2) | 1 (0.1) | 1 (0.5) | 0.31 |
| Rhinorrhea, N (%) | 120 (10.3) | 100 (10.3) | 20 (10.1) | 0.99 |
| Score, median (IQR) | 3 (2, 4) | 3 (2, 4) | 2.5 (1.25, 3.75) | 0.37 |
| As first symptom, N (%) | 9 (0.8) | 8 (0.8) | 1 (0.5) | 0.99 |
| Nasal itching, N (%) | 57 (4.9) | 45 (4.6) | 12 (6.0) | 0.37 |
| Score, median (IQR) | 2 (1.5, 3) | 2 (2, 3.5) | 2 (1, 2) | 0.08 |
| As first symptom, N (%) | 1 (0.1) | 1 (0.1) | 0 (0) | 0.99 |
| Sneezing | 129 (11.0) | 104 (10.7) | 25 (12.6) | 0.46 |
| Score, median (IQR) | 2 (1, 3) | 2 (1, 3) | 2 (1, 2.5) | 0.77 |
| As first symptom, N (%) | 1 (0.1) | 1 (0.1) | 0 (0) | 0.99 |
| Loss of smell | 134 (11.4) | 110 (11.3) | 24 (12.1) | 0.71 |
| Score, median (IQR) | 5 (4, 9) | 5 (4, 8) | 7.5 (3.25, 10) | 0.37 |
| As first symptom, N (%) | 3 (0.3) | 3 (0.3) | 0 (0) | 0.99 |
| Loss of smell recovered<br>, N (%) | 110 (82.1%) | 90 (81.8%) | 20 (83.3%) | 0.99 |
| Recovery time, days, median<br>(IQR) | 8 (6, 13.25) | 8 (5, 12.25) | 8 (6.25, 14) | 0.67 |
| Loss of taste | 242 (20.6) | 199 (20.5) | 43 (21.6) | 0.70 |
| Score, median (IQR) | 6 (4, 8) | 6 (4, 8) | 7 (5, 9) | **0.03** |
| As first symptom, N (%) | 5 (0.4) | 4 (0.4) | 1 (0.5) | 0.99 |
| Loss of taste recovered, N (%) | 231 (95.5%) | 190 (95.5%) | 41 (95.3%) | 0.99 |
| Recovery time, days, median<br>(IQR) | 7 (5, 14) | 7 (5, 14) | 7 (5, 14.5) | 0.76 |

Abbreviations: IQR, interquartile range; AR, allergic rhinitis, CRS, chronic rhinosinusitis; COPD, chronic obstructive pulmonary disease. Data are presented as medians and interquartile ranges (IQR) for continuous variables and numbers with percentage for categorical variables. *P* values were calculated from Kruskal- Wallis, $\chi^2$ test or Fisher's exact test.

Of the 1172 cases, 399 (25.1%) reported with at least one comorbidity, and 17.3% having one or more respiratory comorbidities, including allergic rhinitis (AR, 9.8%), chronic rhinosinusitis (CRS, 6.1%), asthma (2.5%) and chronic obstructive pulmonary disease (COPD, 0.9%). The frequency of patients with at least one nasal symptom was up to 29.2%, including nasal obstruction (8.6%; median score, 3), rhinorrhea (10.3%; median score, 3), nasal itching (4.9%; median score, 2), sneezing (11.0%; median score, 2), loss of smell (11.4%; median score, 5), and loss of taste (20.6%; median score, 6). The incidence of symptom reported as the first onset symptom was < 1% for each individual nasal symptom, including loss of smell and taste. No difference in frequency of patients with loss of smell or taste disorder was found between severe and non-severe COVID-19 cases (Fig 1, A). The scores of loss of taste but not smell were significantly higher in the patients with severe vs. non-severe COVID-19 (7 [5-9] vs. 6 [4-8]; *P* = 0.03) (Fig 1, B). No difference in frequency or score for the other nasal symptoms was found between severe and non-severe disease. The data is shown in Table 1 and Fig 1.

**Figure 1.**
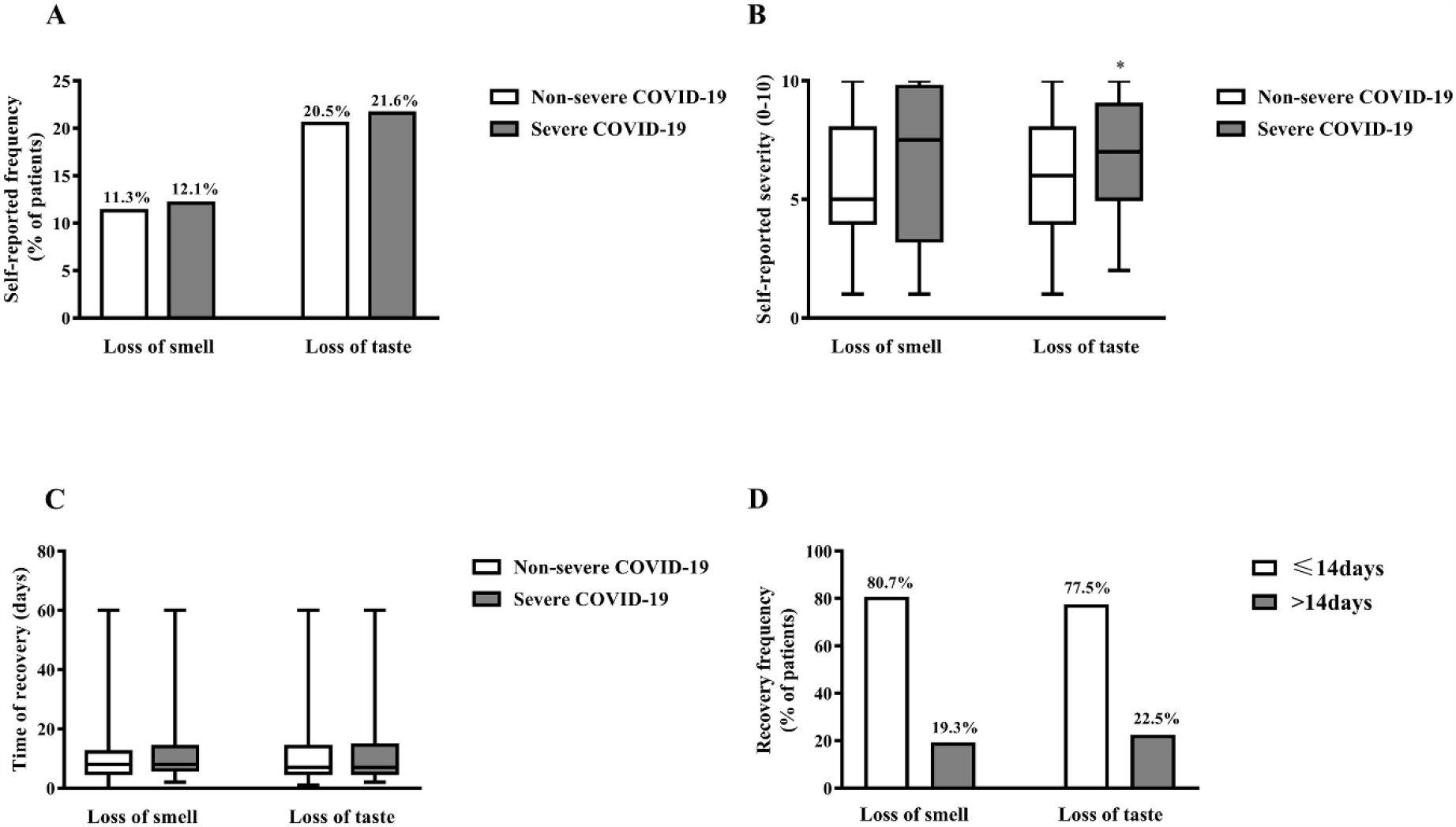
The impact of coronavirus disease 2019 (COVID-19) on smell and taste. (A) The prevalence of self-reported loss of smell and taste in severe and non-severe COVID-19 patients. The frequencies are indicated on the top of the columns. (B) The severity of self-reported loss of smell and taste in severe and non-severe COVID-19 patients. Severity of self-reported loss of smell and taste symptom was scored by patients on a numerical rating scale of 0-10, with 0 being “no complaint whatsoever” and 10 being “the worst imaginable complaint. (C) The recovery time of self-reported smell and state dysfunction in severe and non-severe COVID-19 patients. (D) The pattern of recovery time for patients with self-reported loss of smell and taste. The frequencies are indicated on the tope of columns. * *P* < 0.05 compared to non-severe COVID-19.

Sense of smell and taste are determined by the chemosensory system of the upper respiratory tract, which could be impacted by the nasal dysfunction [22]. Hence, we analyzed the relations between the scores of loss of taste and smell and other nasal symptoms. We failed to find any correlation between loss of taste or loss of smell scores and scores of the other nasal symptoms (Fig E1 in the Online Supplement). However, loss of taste showed mild positive correlation with loss of sense of smell (ρ = 0.25, *P* < 0.01) (Fig E1 in the Online Supplement).

Given the possibility that some patients might not well distinguish the taste and smell disorder [21, 22], we compared the differences among the patients only with one smell or taste disorder, the patients with both smell and taste disorder, and the patients without any of these two symptoms. We found that the patients without loss of smell and taste were significant elder than the patients in the other two groups (62 [48-69] years vs 59 [46-67] years, 57.5 [42.75-66] years, *P* = 0.03). No other difference of clinical characteristics and laboratory measurements was found among the patients in three groups. The data is shown in Table E1 in the Online Supplement.

### The recovery of olfactory and taste function

We found that 82.1% (110/134) of patients with loss of sense of smell and 95.5% (231/242) of patients with loss of taste recovered in one month after discharge. The symptomatic duration days showed no difference between the patients with loss of smell and taste (8 [6-13.25] vs 7 [5-14] days, *P* = 0.52) (Fig 1, C). Most of them recovered in 14 days after onset of symptom (Fig 1, D). No difference in recovery frequency of smell function was found between severe (83.3%) and non-severe cases (81.8%) (*P* = 0.99) (Table 1). As to taste disorder, 95.3% patients with severe COVID-19 and 95.5% patients with non-severe COVID-19 recovered, and no difference showed neither (*P* = 0.99) (Table 1). The data is also shown in Table 1. Due to the limited number of patients with unrecovered symptoms, we did not compare the differences between recovered and unrecovered patients with loss of smell or loss of taste.

### Clinical characteristics of COVID-19 patients with different severity of taste and smell disorder

Since the taste disorder was the most common upper respiratory tract symptoms and showed positive correlation to the symptom of loss of smell, we subsequently compared the differences among the COVID-19 patients with different severity of loss of taste (Table 2) and loss of smell (Table 3). First, we divided 242 COVID-19 cases with loss of taste into mild (score, 1-3; 19.0%), moderate (score, 4-7; 48.8%) and severe (score, 8-10; 32.2%) group by symptom scores, similar to the visual analogue scale system of nasal symptoms [19, 20]. More COVID-19 cases with severe illness were found in the severe loss of taste group than in the moderate loss of taste group (26.9% vs 12.7%, *P* = 0.03). The symptom duration of loss of taste was significantly longer in moderate and severe taste dysfunction group as compared to the mild taste dysfunction group (8 days [6-13.25], 10 days [5-15] vs 5 days [3.5-8], *P* < 0.01) (Fig 2). In addition, serum levels of interleukin-6 (IL-6) and lactate dehydrogenase (LDH) were significantly increased in patients with severe compared to mild and moderate loss of taste group (Table 2). Levels of IL-8 were significantly increased in severe and moderate compared to mild loss of taste group (Table 2). The characteristics of the patients with different severity of taste disorder are shown in Table 2. In addition, IL-6 and LDH showed mild positive correlations to the symptom scores of loss of taste (ρ = 0.15, *P* = 0.03; ρ = 0.21, *P* < 0.01, respectively; Fig 3).

**Table 2.**
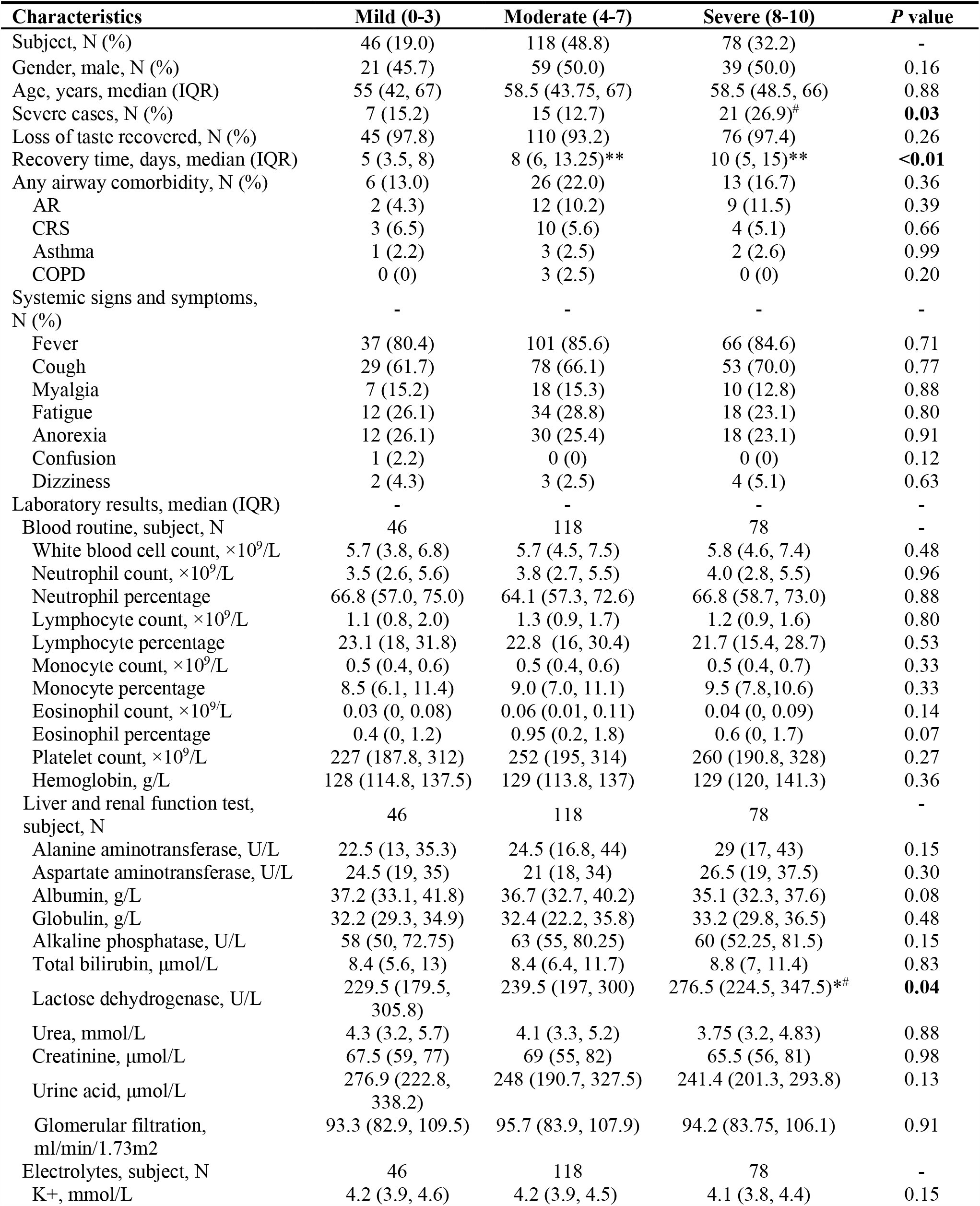
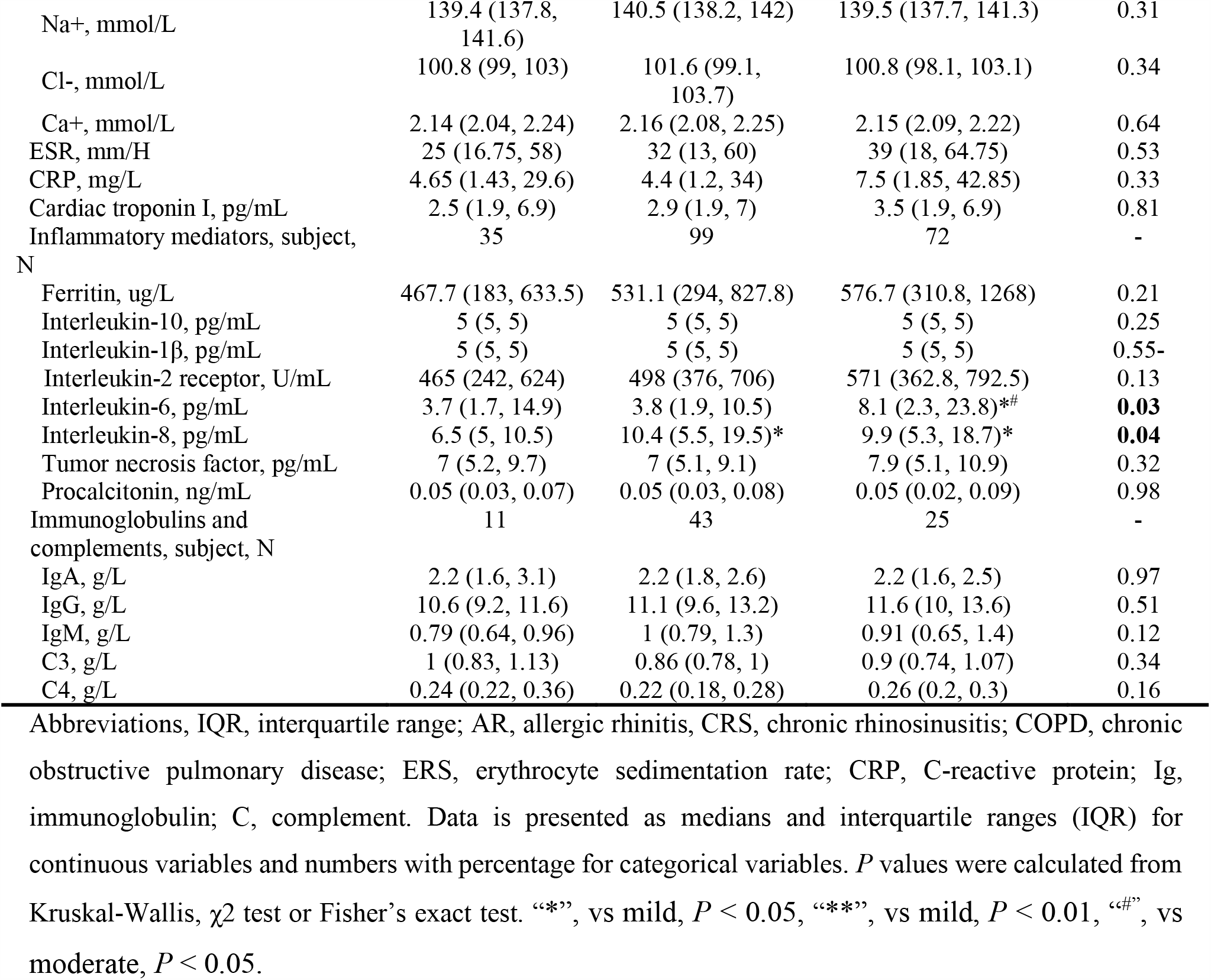
The comparison of 242 patients with different severity of loss of taste.

| Characteristics | Mild (0-3) | Moderate (4-7) | Severe (8-10) | P value |
| --- | --- | --- | --- | --- |
| Subject, N (%) | 46 (19.0) | 118 (48.8) | 78 (32.2) | - |
| Gender, male, N (%) | 21 (45.7) | 59 (50.0) | 39 (50.0) | 0.16 |
| Age, years, median (IQR) | 55 (42, 67) | 58.5 (43.75, 67) | 58.5 (48.5, 66) | 0.88 |
| Severe cases, N (%) | 7 (15.2) | 15 (12.7) | 21 (26.9)<sup>#</sup> | **0.03** |
| Loss of taste recovered, N (%) | 45 (97.8) | 110 (93.2) | 76 (97.4) | 0.26 |
| Recovery time, days, median (IQR) | 5 (3.5, 8) | 8 (6, 13.25)** | 10 (5, 15)** | **<0.01** |
| Any airway comorbidity, N (%) | 6 (13.0) | 26 (22.0) | 13 (16.7) | 0.36 |
| AR | 2 (4.3) | 12 (10.2) | 9 (11.5) | 0.39 |
| CRS | 3 (6.5) | 10 (5.6) | 4 (5.1) | 0.66 |
| Asthma | 1 (2.2) | 3 (2.5) | 2 (2.6) | 0.99 |
| COPD | 0 (0) | 3 (2.5) | 0 (0) | 0.20 |
| Systemic signs and symptoms, N (%) | - | - | - | - |
| Fever | 37 (80.4) | 101 (85.6) | 66 (84.6) | 0.71 |
| Cough | 29 (61.7) | 78 (66.1) | 53 (70.0) | 0.77 |
| Myalgia | 7 (15.2) | 18 (15.3) | 10 (12.8) | 0.88 |
| Fatigue | 12 (26.1) | 34 (28.8) | 18 (23.1) | 0.80 |
| Anorexia | 12 (26.1) | 30 (25.4) | 18 (23.1) | 0.91 |
| Confusion | 1 (2.2) | 0 (0) | 0 (0) | 0.12 |
| Dizziness | 2 (4.3) | 3 (2.5) | 4 (5.1) | 0.63 |
| Laboratory results, median (IQR) | - | - | - | - |
| Blood routine, subject, N | 46 | 118 | 78 | - |
| White blood cell count, $\times 10^9/L$ | 5.7 (3.8, 6.8) | 5.7 (4.5, 7.5) | 5.8 (4.6, 7.4) | 0.48 |
| Neutrophil count, $\times 10^9/L$ | 3.5 (2.6, 5.6) | 3.8 (2.7, 5.5) | 4.0 (2.8, 5.5) | 0.96 |
| Neutrophil percentage | 66.8 (57.0, 75.0) | 64.1 (57.3, 72.6) | 66.8 (58.7, 73.0) | 0.88 |
| Lymphocyte count, $\times 10^9/L$ | 1.1 (0.8, 2.0) | 1.3 (0.9, 1.7) | 1.2 (0.9, 1.6) | 0.80 |
| Lymphocyte percentage | 23.1 (18, 31.8) | 22.8 (16, 30.4) | 21.7 (15.4, 28.7) | 0.53 |
| Monocyte count, $\times 10^9/L$ | 0.5 (0.4, 0.6) | 0.5 (0.4, 0.6) | 0.5 (0.4, 0.7) | 0.33 |
| Monocyte percentage | 8.5 (6.1, 11.4) | 9.0 (7.0, 11.1) | 9.5 (7.8, 10.6) | 0.33 |
| Eosinophil count, $\times 10^9/L$ | 0.03 (0, 0.08) | 0.06 (0.01, 0.11) | 0.04 (0, 0.09) | 0.14 |
| Eosinophil percentage | 0.4 (0, 1.2) | 0.95 (0.2, 1.8) | 0.6 (0, 1.7) | 0.07 |
| Platelet count, $\times 10^9/L$ | 227 (187.8, 312) | 252 (195, 314) | 260 (190.8, 328) | 0.27 |
| Hemoglobin, g/L | 128 (114.8, 137.5) | 129 (113.8, 137) | 129 (120, 141.3) | 0.36 |
| Liver and renal function test, subject, N | 46 | 118 | 78 | - |
| Alanine aminotransferase, U/L | 22.5 (13, 35.3) | 24.5 (16.8, 44) | 29 (17, 43) | 0.15 |
| Aspartate aminotransferase, U/L | 24.5 (19, 35) | 21 (18, 34) | 26.5 (19, 37.5) | 0.30 |
| Albumin, g/L | 37.2 (33.1, 41.8) | 36.7 (32.7, 40.2) | 35.1 (32.3, 37.6) | 0.08 |
| Globulin, g/L | 32.2 (29.3, 34.9) | 32.4 (22.2, 35.8) | 33.2 (29.8, 36.5) | 0.48 |
| Alkaline phosphatase, U/L | 58 (50, 72.75) | 63 (55, 80.25) | 60 (52.25, 81.5) | 0.15 |
| Total bilirubin, $\mu\text{mol/L}$ | 8.4 (5.6, 13) | 8.4 (6.4, 11.7) | 8.8 (7, 11.4) | 0.83 |
| Lactose dehydrogenase, U/L | 229.5 (179.5, 305.8) | 239.5 (197, 300) | 276.5 (224.5, 347.5)*<sup>#</sup> | **0.04** |
| Urea, mmol/L | 4.3 (3.2, 5.7) | 4.1 (3.3, 5.2) | 3.75 (3.2, 4.83) | 0.88 |
| Creatinine, $\mu\text{mol/L}$ | 67.5 (59, 77) | 69 (55, 82) | 65.5 (56, 81) | 0.98 |
| Urine acid, $\mu\text{mol/L}$ | 276.9 (222.8, 338.2) | 248 (190.7, 327.5) | 241.4 (201.3, 293.8) | 0.13 |
| Glomerular filtration, ml/min/1.73m<sup>2</sup> | 93.3 (82.9, 109.5) | 95.7 (83.9, 107.9) | 94.2 (83.75, 106.1) | 0.91 |
| Electrolytes, subject, N | 46 | 118 | 78 | - |
| K<sup>+</sup>, mmol/L | 4.2 (3.9, 4.6) | 4.2 (3.9, 4.5) | 4.1 (3.8, 4.4) | 0.15 |
| Na<sup>+</sup>, mmol/L | 139.4 (137.8, 141.6) | 140.5 (138.2, 142) | 139.5 (137.7, 141.3) | 0.31 |
| Cl<sup>-</sup>, mmol/L | 100.8 (99, 103) | 101.6 (99.1, 103.7) | 100.8 (98.1, 103.1) | 0.34 |
| Ca<sup>+</sup>, mmol/L | 2.14 (2.04, 2.24) | 2.16 (2.08, 2.25) | 2.15 (2.09, 2.22) | 0.64 |
| ESR, mm/H | 25 (16.75, 58) | 32 (13, 60) | 39 (18, 64.75) | 0.53 |
| CRP, mg/L | 4.65 (1.43, 29.6) | 4.4 (1.2, 34) | 7.5 (1.85, 42.85) | 0.33 |
| Cardiac troponin I, pg/mL | 2.5 (1.9, 6.9) | 2.9 (1.9, 7) | 3.5 (1.9, 6.9) | 0.81 |
| Inflammatory mediators, subject, N | 35 | 99 | 72 | - |
| Ferritin, ug/L | 467.7 (183, 633.5) | 531.1 (294, 827.8) | 576.7 (310.8, 1268) | 0.21 |
| Interleukin-10, pg/mL | 5 (5, 5) | 5 (5, 5) | 5 (5, 5) | 0.25 |
| Interleukin-1β, pg/mL | 5 (5, 5) | 5 (5, 5) | 5 (5, 5) | 0.55- |
| Interleukin-2 receptor, U/mL | 465 (242, 624) | 498 (376, 706) | 571 (362.8, 792.5) | 0.13 |
| Interleukin-6, pg/mL | 3.7 (1.7, 14.9) | 3.8 (1.9, 10.5) | 8.1 (2.3, 23.8)*# | **0.03** |
| Interleukin-8, pg/mL | 6.5 (5, 10.5) | 10.4 (5.5, 19.5)* | 9.9 (5.3, 18.7)* | **0.04** |
| Tumor necrosis factor, pg/mL | 7 (5.2, 9.7) | 7 (5.1, 9.1) | 7.9 (5.1, 10.9) | 0.32 |
| Procalcitonin, ng/mL | 0.05 (0.03, 0.07) | 0.05 (0.03, 0.08) | 0.05 (0.02, 0.09) | 0.98 |
| Immunoglobulins and complements, subject, N | 11 | 43 | 25 | - |
| IgA, g/L | 2.2 (1.6, 3.1) | 2.2 (1.8, 2.6) | 2.2 (1.6, 2.5) | 0.97 |
| IgG, g/L | 10.6 (9.2, 11.6) | 11.1 (9.6, 13.2) | 11.6 (10, 13.6) | 0.51 |
| IgM, g/L | 0.79 (0.64, 0.96) | 1 (0.79, 1.3) | 0.91 (0.65, 1.4) | 0.12 |
| C3, g/L | 1 (0.83, 1.13) | 0.86 (0.78, 1) | 0.9 (0.74, 1.07) | 0.34 |
| C4, g/L | 0.24 (0.22, 0.36) | 0.22 (0.18, 0.28) | 0.26 (0.2, 0.3) | 0.16 |

Abbreviations, IQR, interquartile range; AR, allergic rhinitis, CRS, chronic rhinosinusitis; COPD, chronic obstructive pulmonary disease; ERS, erythrocyte sedimentation rate; CRP, C-reactive protein; Ig, immunoglobulin; C, complement. Data is presented as medians and interquartile ranges (IQR) for continuous variables and numbers with percentage for categorical variables. *P* values were calculated from Kruskal-Wallis, $\chi^2$ test or Fisher's exact test. “\*”, vs mild, *P* < 0.05, “\*\*\*”, vs mild, *P* < 0.01, “#”, vs moderate, *P* < 0.05.

**Table 3.**
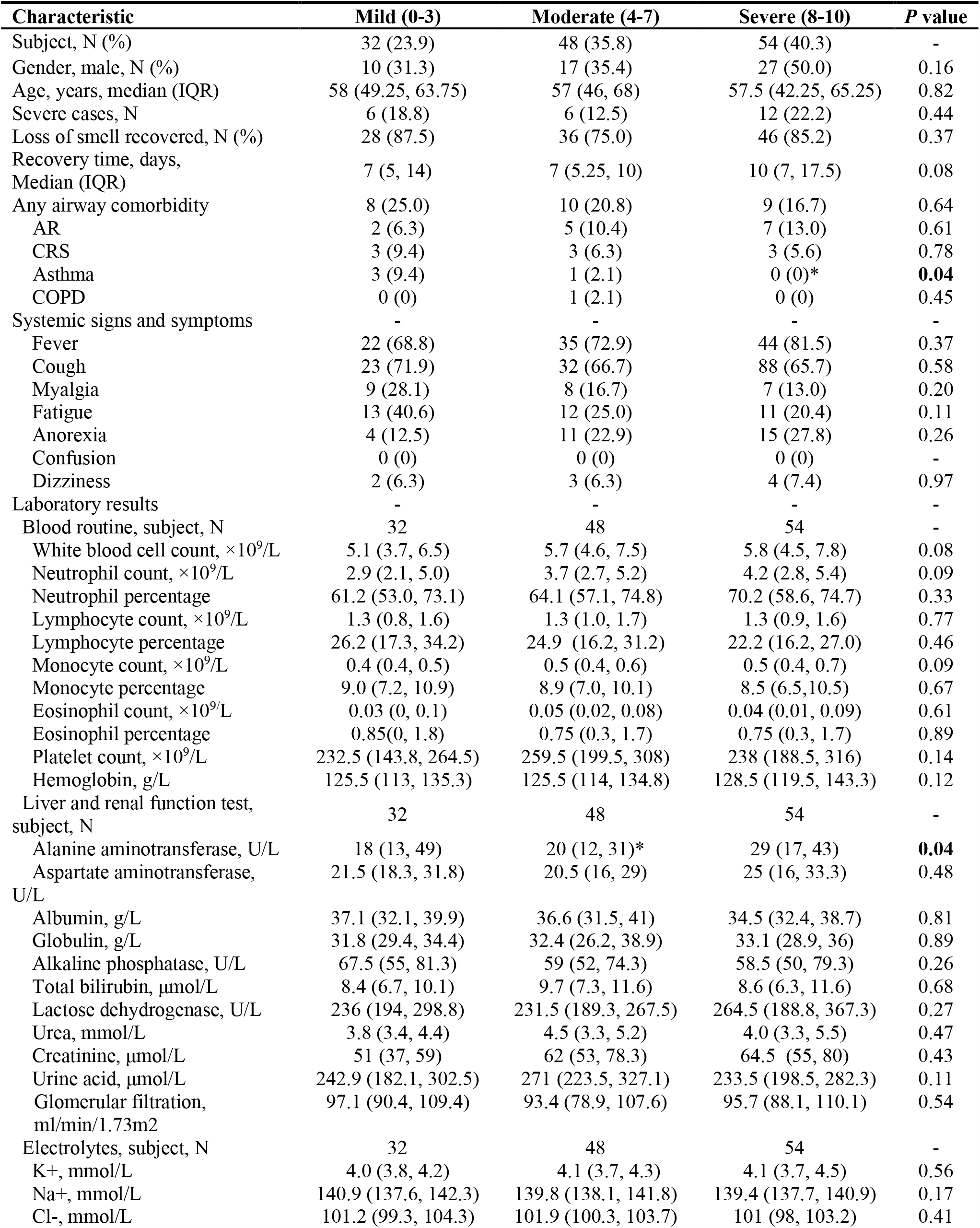
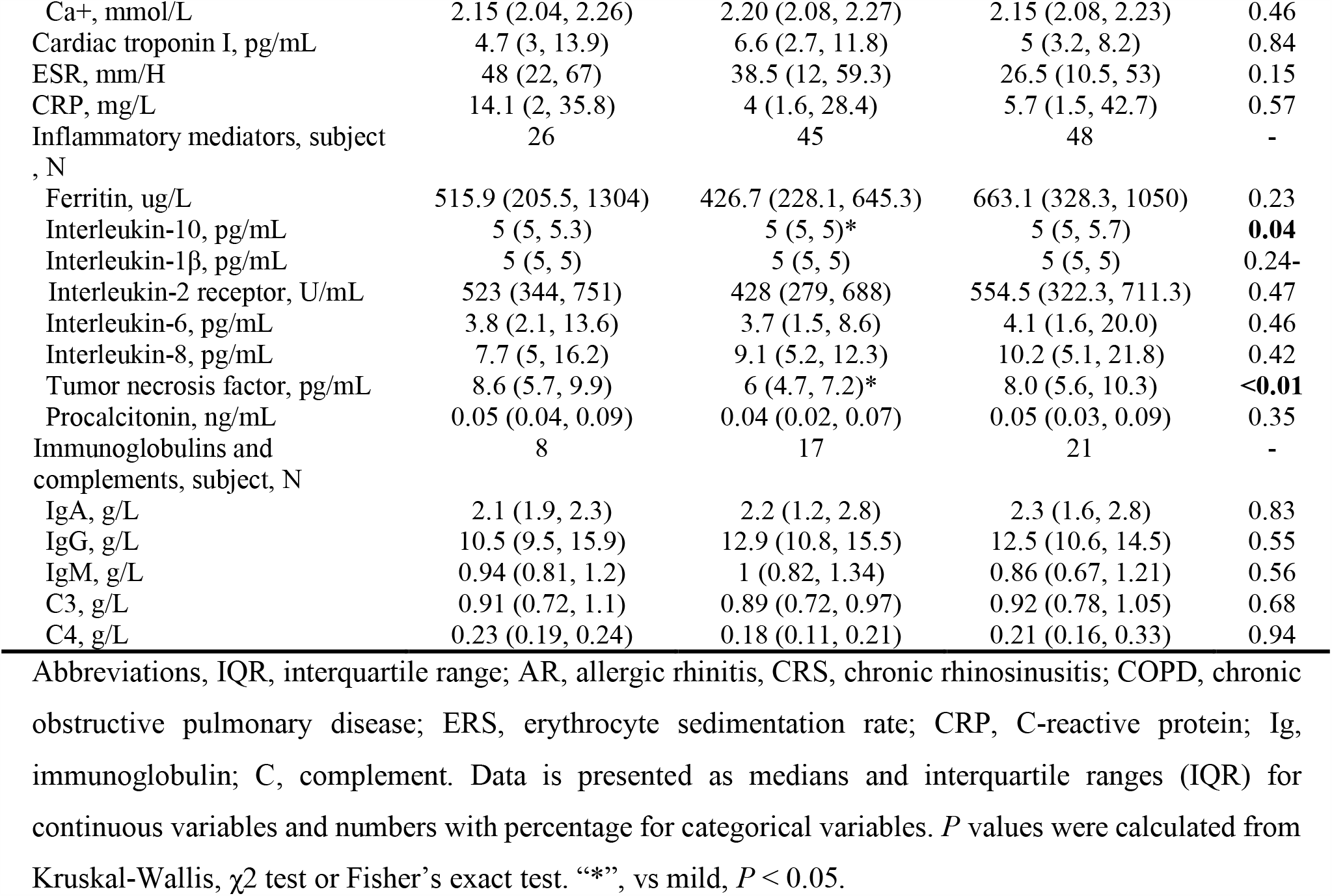
The comparison of 134 patients with different severity of loss of smell.

| Characteristic | Mild (0-3) | Moderate (4-7) | Severe (8-10) | P value |
| --- | --- | --- | --- | --- |
| Subject, N (%) | 32 (23.9) | 48 (35.8) | 54 (40.3) | - |
| Gender, male, N (%) | 10 (31.3) | 17 (35.4) | 27 (50.0) | 0.16 |
| Age, years, median (IQR) | 58 (49.25, 63.75) | 57 (46, 68) | 57.5 (42.25, 65.25) | 0.82 |
| Severe cases, N | 6 (18.8) | 6 (12.5) | 12 (22.2) | 0.44 |
| Loss of smell recovered, N (%) | 28 (87.5) | 36 (75.0) | 46 (85.2) | 0.37 |
| Recovery time, days,<br>Median (IQR) | 7 (5, 14) | 7 (5.25, 10) | 10 (7, 17.5) | 0.08 |
| Any airway comorbidity | 8 (25.0) | 10 (20.8) | 9 (16.7) | 0.64 |
| AR | 2 (6.3) | 5 (10.4) | 7 (13.0) | 0.61 |
| CRS | 3 (9.4) | 3 (6.3) | 3 (5.6) | 0.78 |
| Asthma | 3 (9.4) | 1 (2.1) | 0 (0)* | **0.04** |
| COPD | 0 (0) | 1 (2.1) | 0 (0) | 0.45 |
| Systemic signs and symptoms | - | - | - | - |
| Fever | 22 (68.8) | 35 (72.9) | 44 (81.5) | 0.37 |
| Cough | 23 (71.9) | 32 (66.7) | 88 (65.7) | 0.58 |
| Myalgia | 9 (28.1) | 8 (16.7) | 7 (13.0) | 0.20 |
| Fatigue | 13 (40.6) | 12 (25.0) | 11 (20.4) | 0.11 |
| Anorexia | 4 (12.5) | 11 (22.9) | 15 (27.8) | 0.26 |
| Confusion | 0 (0) | 0 (0) | 0 (0) | - |
| Dizziness | 2 (6.3) | 3 (6.3) | 4 (7.4) | 0.97 |
| Laboratory results | - | - | - | - |
| Blood routine, subject, N | 32 | 48 | 54 | - |
| White blood cell count, $\times 10^9/L$ | 5.1 (3.7, 6.5) | 5.7 (4.6, 7.5) | 5.8 (4.5, 7.8) | 0.08 |
| Neutrophil count, $\times 10^9/L$ | 2.9 (2.1, 5.0) | 3.7 (2.7, 5.2) | 4.2 (2.8, 5.4) | 0.09 |
| Neutrophil percentage | 61.2 (53.0, 73.1) | 64.1 (57.1, 74.8) | 70.2 (58.6, 74.7) | 0.33 |
| Lymphocyte count, $\times 10^9/L$ | 1.3 (0.8, 1.6) | 1.3 (1.0, 1.7) | 1.3 (0.9, 1.6) | 0.77 |
| Lymphocyte percentage | 26.2 (17.3, 34.2) | 24.9 (16.2, 31.2) | 22.2 (16.2, 27.0) | 0.46 |
| Monocyte count, $\times 10^9/L$ | 0.4 (0.4, 0.5) | 0.5 (0.4, 0.6) | 0.5 (0.4, 0.7) | 0.09 |
| Monocyte percentage | 9.0 (7.2, 10.9) | 8.9 (7.0, 10.1) | 8.5 (6.5, 10.5) | 0.67 |
| Eosinophil count, $\times 10^9/L$ | 0.03 (0, 0.1) | 0.05 (0.02, 0.08) | 0.04 (0.01, 0.09) | 0.61 |
| Eosinophil percentage | 0.85 (0, 1.8) | 0.75 (0.3, 1.7) | 0.75 (0.3, 1.7) | 0.89 |
| Platelet count, $\times 10^9/L$ | 232.5 (143.8, 264.5) | 259.5 (199.5, 308) | 238 (188.5, 316) | 0.14 |
| Hemoglobin, g/L | 125.5 (113, 135.3) | 125.5 (114, 134.8) | 128.5 (119.5, 143.3) | 0.12 |
| Liver and renal function test,<br>subject, N | 32 | 48 | 54 | - |
| Alanine aminotransferase, U/L | 18 (13, 49) | 20 (12, 31)* | 29 (17, 43) | **0.04** |
| Aspartate aminotransferase,<br>U/L | 21.5 (18.3, 31.8) | 20.5 (16, 29) | 25 (16, 33.3) | 0.48 |
| Albumin, g/L | 37.1 (32.1, 39.9) | 36.6 (31.5, 41) | 34.5 (32.4, 38.7) | 0.81 |
| Globulin, g/L | 31.8 (29.4, 34.4) | 32.4 (26.2, 38.9) | 33.1 (28.9, 36) | 0.89 |
| Alkaline phosphatase, U/L | 67.5 (55, 81.3) | 59 (52, 74.3) | 58.5 (50, 79.3) | 0.26 |
| Total bilirubin, $\mu\text{mol/L}$ | 8.4 (6.7, 10.1) | 9.7 (7.3, 11.6) | 8.6 (6.3, 11.6) | 0.68 |
| Lactose dehydrogenase, U/L | 236 (194, 298.8) | 231.5 (189.3, 267.5) | 264.5 (188.8, 367.3) | 0.27 |
| Urea, mmol/L | 3.8 (3.4, 4.4) | 4.5 (3.3, 5.2) | 4.0 (3.3, 5.5) | 0.47 |
| Creatinine, $\mu\text{mol/L}$ | 51 (37, 59) | 62 (53, 78.3) | 64.5 (55, 80) | 0.43 |
| Urine acid, $\mu\text{mol/L}$ | 242.9 (182.1, 302.5) | 271 (223.5, 327.1) | 233.5 (198.5, 282.3) | 0.11 |
| Glomerular filtration,<br>ml/min/1.73m<sup>2</sup> | 97.1 (90.4, 109.4) | 93.4 (78.9, 107.6) | 95.7 (88.1, 110.1) | 0.54 |
| Electrolytes, subject, N | 32 | 48 | 54 | - |
| K<sup>+</sup>, mmol/L | 4.0 (3.8, 4.2) | 4.1 (3.7, 4.3) | 4.1 (3.7, 4.5) | 0.56 |
| Na<sup>+</sup>, mmol/L | 140.9 (137.6, 142.3) | 139.8 (138.1, 141.8) | 139.4 (137.7, 140.9) | 0.17 |
| Cl<sup>-</sup>, mmol/L | 101.2 (99.3, 104.3) | 101.9 (100.3, 103.7) | 101 (98, 103.2) | 0.41 |
| Ca<sup>+</sup>, mmol/L | 2.15 (2.04, 2.26) | 2.20 (2.08, 2.27) | 2.15 (2.08, 2.23) | 0.46 |
| Cardiac troponin I, pg/mL | 4.7 (3, 13.9) | 6.6 (2.7, 11.8) | 5 (3.2, 8.2) | 0.84 |
| ESR, mm/H | 48 (22, 67) | 38.5 (12, 59.3) | 26.5 (10.5, 53) | 0.15 |
| CRP, mg/L | 14.1 (2, 35.8) | 4 (1.6, 28.4) | 5.7 (1.5, 42.7) | 0.57 |
| Inflammatory mediators, subject, N | 26 | 45 | 48 | - |
| Ferritin, ug/L | 515.9 (205.5, 1304) | 426.7 (228.1, 645.3) | 663.1 (328.3, 1050) | 0.23 |
| Interleukin-10, pg/mL | 5 (5, 5.3) | 5 (5, 5)* | 5 (5, 5.7) | **0.04** |
| Interleukin-1β, pg/mL | 5 (5, 5) | 5 (5, 5) | 5 (5, 5) | 0.24- |
| Interleukin-2 receptor, U/mL | 523 (344, 751) | 428 (279, 688) | 554.5 (322.3, 711.3) | 0.47 |
| Interleukin-6, pg/mL | 3.8 (2.1, 13.6) | 3.7 (1.5, 8.6) | 4.1 (1.6, 20.0) | 0.46 |
| Interleukin-8, pg/mL | 7.7 (5, 16.2) | 9.1 (5.2, 12.3) | 10.2 (5.1, 21.8) | 0.42 |
| Tumor necrosis factor, pg/mL | 8.6 (5.7, 9.9) | 6 (4.7, 7.2)* | 8.0 (5.6, 10.3) | **<0.01** |
| Procalcitonin, ng/mL | 0.05 (0.04, 0.09) | 0.04 (0.02, 0.07) | 0.05 (0.03, 0.09) | 0.35 |
| Immunoglobulins and complements, subject, N | 8 | 17 | 21 | - |
| IgA, g/L | 2.1 (1.9, 2.3) | 2.2 (1.2, 2.8) | 2.3 (1.6, 2.8) | 0.83 |
| IgG, g/L | 10.5 (9.5, 15.9) | 12.9 (10.8, 15.5) | 12.5 (10.6, 14.5) | 0.55 |
| IgM, g/L | 0.94 (0.81, 1.2) | 1 (0.82, 1.34) | 0.86 (0.67, 1.21) | 0.56 |
| C3, g/L | 0.91 (0.72, 1.1) | 0.89 (0.72, 0.97) | 0.92 (0.78, 1.05) | 0.68 |
| C4, g/L | 0.23 (0.19, 0.24) | 0.18 (0.11, 0.21) | 0.21 (0.16, 0.33) | 0.94 |

Abbreviations, IQR, interquartile range; AR, allergic rhinitis, CRS, chronic rhinosinusitis; COPD, chronic obstructive pulmonary disease; ERS, erythrocyte sedimentation rate; CRP, C-reactive protein; Ig, immunoglobulin; C, complement. Data is presented as medians and interquartile ranges (IQR) for continuous variables and numbers with percentage for categorical variables. *P* values were calculated from Kruskal-Wallis, $\chi^2$ test or Fisher's exact test. “\*”, vs mild, *P* < 0.05.

**Figure 2.**
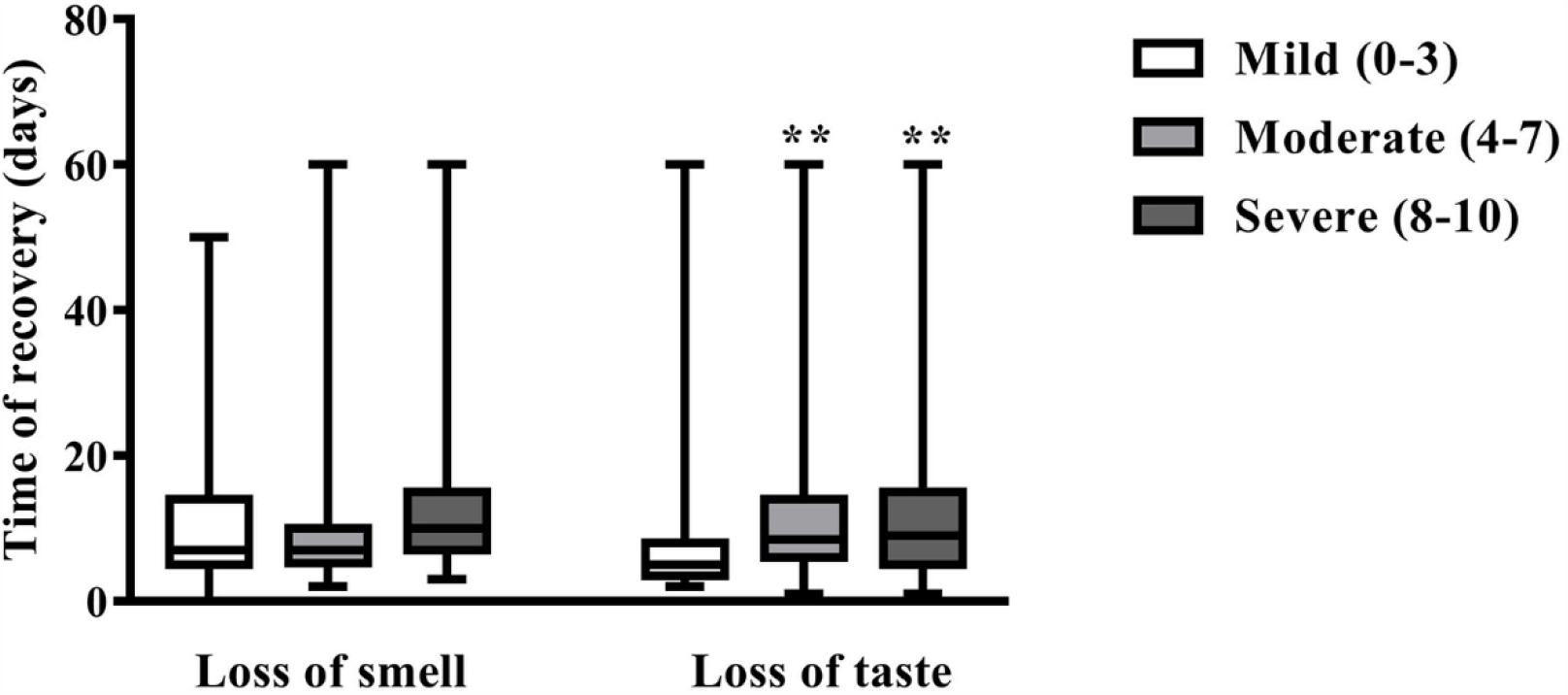
The recovery time of patients with different severity of loss of smell and loss of taste. Severity of self-reported loss of smell and taste symptom was scored by patients on a numerical rating scale of 0-10, with 0 being “no complaint whatsoever” and 10 being “the worst imaginable complaint. ** *P* < 0.01 compared to mild loss of taste.

**Figure 3.**
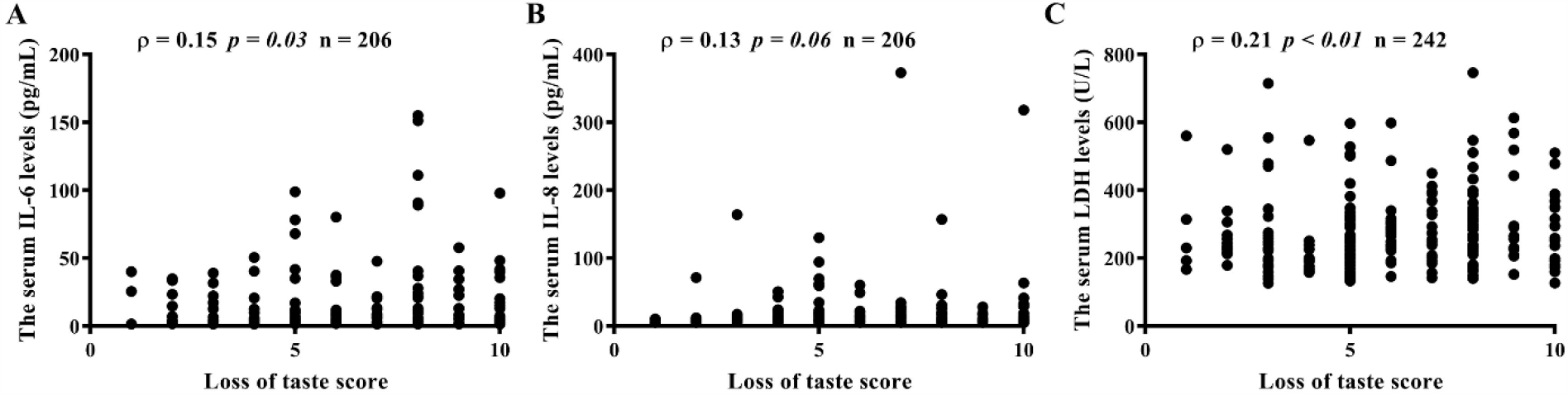
Correlations between loss of taste scores and serum levels of IL-6 (A), IL-8 (B), lactose dehydrogenase (LDH) (C). Severity of self-reported loss of taste was scored by patients on a numerical rating scale of 0-10, with 0 being “no complaint whatsoever” and 10 being “the worst imaginable complaint.

As to the comparison among different severity of loss of smell, we divided the 134 patients with loss of sense of smell into mild (score, 1-3, 23.9%), moderate (score, 4-7; 35.8%) and severe (score, 8-10; 40.3%) group. No prominent difference in clinical and laboratory measurements was found among the patients with different severity (Table 3 and Fig 2).

## Discussion

To the best of our knowledge, this is the first study on detailed information of manifestations of the upper respiratory tract of the hospitalized patients with COVID-19 in China. We reported the manifestations of upper respiratory tract in a cohort of 1,172 patients with laboratory-confirmed COVID-19 in Wuhan, China. Overall, 17% subjects were categorized as severe COVID-19, this percentage being similar with previous reports from China [7, 10]. Most of the common systemic signs and symptoms of disease such as fever, fatigue, and anorexia, etc. reported in previous studies were also observed in our cohort [2, 7, 10]. Nevertheless, under urgent admission of COVID-19 patients, compared to systemic symptoms and comorbidities, nasal symptoms and upper airway comorbidities were very likely incompletely documented and underestimated in previous studies based on medical record analysis [7, 23]. In order to overcome this limitation in real-world setting, we did recalled questionnaires by phone call to reevaluate the presence of airway comorbidities, and frequency and severity of nasal symptoms. As a result, the frequencies of COVID-19 patients with AR and CRS were 9.8% and 6.1%, respectively. Previous studies have reported that adult AR and CRS in general population in China were 17.6% and 8%, respectively, these being higher than the reported for COVID-19 [24, 25]. Recent studies indicate that the increasing eosinophils may be an indicator of COVID-19 improvement [26, 27]. Hence, we cannot rule out the possibility that the comorbidities of allergic and eosinophilic diseases such as AR might be potential protective factors for severe COVID-19.

Here, we found that upper respiratory tract symptoms were identified in 29.2% patients with COVID-19, with nasal obstruction, rhinorrhea, nasal itching and sneezing presenting in mild/moderate severity and loss of smell and taste presenting in moderate/severe severity. All of these upper respiratory tract symptoms have been commonly reported in other respiratory viral infection, such as influenza and rhinovirus [28-30]. Our data suggest that similar with the common respiratory virtues, SARS-CoV-2 may be able to infect upper respiratory tract mucosa and cause similar symptoms [22, 29, 31]. In addition, the result in our Wuhan cohort revealed that the self-reported symptoms of upper respiratory tract were not the first symptom in most COVID-19 patients, doubting the diagnostic value of these symptoms preceding the onset of full-blown clinical disease. However, our study only included hospitalized COVID-19 patients while the clinical manifestations of COVID-19 can range from asymptomatic infection to severe pneumonia [2, 7, 32]. Further investigations on non-hospitalized infected patients and patients with likely sudden onset anosmia will help to determine the role of upper airway symptoms, particular loss of smell and taste, as a screening tool for COVID-19.

Interestingly, the loss of taste and smell were the most common upper respiratory tract symptoms observed in COVID-19 patients in Wuhan, with 17.5% patients having only smell or taste disorder and 7.3% patients having both symptoms. By integrating medical record analysis and follow-up confirmation and reevaluation after discharge, we reported higher rates of smell and taste disorder in the hospitalized patients than an early report from Wuhan, which relied on medical record analysis only and had a smaller sample size [17]. However, the rates of smell and taste disorder in our cohort were much lower than those reported in Europe and America [13-16, 33]. It may be related to the different characteristics of COVID-19 patients. In our study, asymptomatic patients and non-hospitalized patients with mild symptoms were not included. It is also likely that people with distinct ethnic/culture background may have different response to SARS-CoV-2 infection. Previously, no large-sample-size investigation of olfactory and taste disorder in the disease caused by either SARS-CoV-1 or middle east respiratory syndrome coronavirus, that belongs to the same family of coronaviruses, was undertaken. A case report revealed the olfactory disorder in a patient infected with SARS-CoV-1 [34]. Nevertheless, olfactory and taste disorders are well known to be widely associated with a number of viral infections [35, 36]. The frequency of patients with loss of smell in the present study was similar to that of post-viral olfactory dysfunction (PVOD) caused by the common respiratory virus, such as influenza with a ratio of about 17% [29, 31]. PVOD can be caused by mechanical obstruction of odorant transmission due to edema of nasal mucosa, and/or the inflammatory impairment of the olfactory neuroepithelium and even central nervous systems [22, 31, 37, 38]. We found that the scores of taste and smell dysfunction showed no correlation with the scores of the other nasal symptoms including nasal obstruction, disfavoring a role of mechanical obstruction in the dysfunction of smell and taste. In fact, the symptom scores indicate that COVID-19 patients suffer from mild/moderate nasal obstruction and rhinorrhea, despite moderate/severe olfactory and taste disorder. SARS-CoV-1 has demonstrated a transneural penetration through the olfactory bulb in mice model [39]. Angiotensin converting enzyme 2, which is used by SARS-CoV-1 and SARS-CoV-2 to invade the host cells, is widely expressed on the epithelial cells in nasal and oral cavity [40, 41]. These evidence suggest a neurological involvement in smell and taste disorder caused by SARS-CoV-2 infection. In this study, COVID-19 patients with severe illness had more severe taste disorder. In addition, serum levels of IL-6, a pro-inflammatory cytokine, were elevated in patients with severe taste disorder, and positively correlated with the loss of taste scores. Previous studies demonstrated that the pro-inflammatory cytokines were able to impair the function of taste buds directly, thus leading to taste dysfunction [42, 43].

The present study revealed that 95.5% patients with loss of taste and 82.1% patients with loss of smell recovered spontaneously, and most of them in 2-week time, although the severe impairment of olfactory and smell function may lead to delayed recovery. The spontaneous recovery may be a result of regeneration of the damaged olfactory epithelium and taste buds [44]. Previously, oral and topical corticosteroids have been proposed to treat viral-associated olfactory loss. However, systemic corticosteroids may impair the viral clearance. Given to the high rate of spontaneous recovery of olfactory and taste function, it seems that there is no need to use corticosteroids for treating olfactory and smell disorder in patients with COVID-19, although their administration can be continued to treat comorbidities such as AR and CRS [45].

We have to acknowledge that there are several limitations of this study. First, the self-reported and recalled symptoms and comorbidities without diagnostic testing, especially for the symptomatic information of loss of smell and taste obtained over a month after onset, might contribute to under- or over-estimation of the prevalence of these symptoms, and the strength of association with the clinical outcomes. Second, the self-report and questionnaire-based evaluation could not clearly discriminate the symptoms of loss of smell and taste in patients with COVID-19. It is the inherent limitation of the study based on self-reported data. Three, it was impossible to include the fatal cases due to the incomplete medical records regarding upper airway symptoms and comorbidities, and impossible telephone recall. Fourth, asymptomatic patients and non-hospitalized patients with mild symptoms were missed in this study. Fifth, we cannot preclude the influence of AR and CRS comorbidity on the presentation of upper airway symptoms during COVID-19. However, most of our patients claimed that their primary AR or CRS were under control at baseline and we found no difference in upper airway symptoms between patients with and without AR or CRS comorbidity (data not shown).

## Conclusion

To the best of our knowledge, this is the largest retrospective cohort study focusing on the symptoms of upper respiratory tract in patients with COVID-19 in China. We found that nearly one third of COVID-19 patients got the upper respiratory tract symptoms. Loss of smell affected one out of ten patients with COVID-19 while loss of taste was the most common upper respiratory tract symptom affecting one out of five patients, being associated with the severity of COVID-19 infection. Finally, about 80% of patients spontaneously recovered from olfactory and taste dysfunction in a short term of 2 weeks.

## Data Availability

Statement by the authors: all data in the manuscript and note links below is available.

https://pan.baidu.com/s/1GeBjkdG1ExzdQIeqk3XvPg

## Notes

### Competing Interest Statement

The authors have declared no competing interest.

### Funding Statement

This study was supported by Natural Science Foundation of Hubei Province of China grant [2018CFB602 (M.Z.)], National Key R&D Program of China (2018YFC0116800), and National Natural Science Foundation of China (NSFC) grants [81630024 and 81920108011 (Z.L.), and 81900925 (J.S.)].

### Author Declarations

This study was approved by the ethics committee of Tongji Hospital and informed consent was obtained from each subject.

